# Ketamine treatment modulates habenular and nucleus accumbens static and dynamic functional connectivity in major depression

**DOI:** 10.1101/2023.12.01.23299282

**Authors:** Brandon Taraku, Joana R. Loureiro, Ashish K. Sahib, Artemis Zavaliangos-Petropulu, Noor Al-Sharif, Amber Leaver, Benjamin Wade, Shantanu Joshi, Roger P. Woods, Randall Espinoza, Katherine L. Narr

## Abstract

Dysfunctional reward processing in major depressive disorder (MDD) involves functional circuitry of the habenula (Hb) and nucleus accumbens (NAc). Ketamine elicits rapid antidepressant and alleviates anhedonia in MDD. To clarify how ketamine perturbs reward circuitry in MDD, we examined how serial ketamine infusions (SKI) modulate static and dynamic functional connectivity (FC) in Hb and NAc networks. MDD participants (n=58, mean age=40.7 years, female=28) received four ketamine infusions (0.5mg/kg) 2-3 times weekly. Resting-state fMRI scans and clinical assessments were collected at baseline and 24 hours post-SKI completion. Static FC (sFC) and dynamic FC variability (dFCv) were calculated from left and right Hb and NAc seeds to all other brain regions. Paired t-tests examined changes in FC pre-to-post SKI, and correlations were used to determine relationships between FC changes with mood and anhedonia. Following SKI, significant increases in left Hb-bilateral visual cortex FC, decreases in left Hb-left inferior parietal cortex FC, and decreases in left NAc-right cerebellum FC occurred. Decreased dFCv between left Hb and right precuneus and visual cortex, and decreased dFCv between right NAc and right visual cortex both significantly correlated with improvements in Hamilton Depression Rating Scale. Decreased FC between left Hb and bilateral visual/parietal cortices as well as increased FC between left NAc and right visual/parietal cortices both significantly correlated with improvements in anhedonia. Subanesthetic ketamine modulates functional pathways linking the Hb and NAc with visual, parietal, and cerebellar regions. Overlapping effects between Hb and NAc functional systems were associated with ketamine’s therapeutic response.

## Introduction

Major depression, characterized by low mood and a loss of pleasure and interest in activities (anhedonia) amongst other symptoms, affects at least 5% of adults worldwide^1^. Though treatable, only one third of patients remit after receiving first-line antidepressants that take weeks to months to induce clinical benefits. About 30-40% of patients still fail to remit^2,3^ after multiple treatment trials and are characterized as having treatment resistant depression (TRD). Anhedonia, a core symptom of depression, has been linked with deficits in reward processing regions of the brain^4,5^. Even amongst patients who remit or respond to standard antidepressants, anhedonic symptoms often persist^6,7^. Anhedonia and reward processing deficits are shown to predict treatment resistance^8,9^, risk of suicide^10–12^ and negatively impact quality of life in people with depression^13,14^.

Ketamine acts as a non-competitive N-methyl-D-aspartate (glutamate) receptor (NMDAR) antagonist that is widely used as an anesthetic in medicine. More recently, subanesthetic ketamine has been shown to rapidly reduce depressive and suicidal symptoms in patients with TRD, though treatment response is typically transient (∼1 week)^15–18^. In contrast to monoaminergic antidepressants, ketamine also effectively reduces symptoms of anhedonia where benefits appear to outlast and be independent of overall antidepressant outcomes^19–22^. Moreover, ketamine treatment is shown to result in improvements in anticipatory, consummatory, and motivation-related reward processes that are considered dimensions of anhedonia in humans and in animal models^19^.

Though the molecular mechanisms of ketamine’s antidepressant action are not yet resolved, available data supports that NMDAR blockade and potentially NMDA independent mechanisms lead to the subsequent activation of downstream signaling pathways to induce rapid dendritic and synaptic remodeling^16,17,23^. These processes of neural plasticity are assumed to then evolve to higher brain systems-level reorganization to influence behavior, including modulation of downstream monoaminergic neurotransmitter systems targeted by standard antidepressants^24–28^.

Preclinical studies suggest that ketamine’s therapeutic effects are at least partially mediated by molecular/cellular changes within the mesolimbic system^29^. The mesolimbic pathway, connecting the midbrain and ventral tegmental area (VTA) with the ventral striatum/nucleus accumbens (NAc) and other limbic and cortical areas, is repeatedly shown to drive reward-related behavior^4,5,30,31^. The NAc, a key node of dopaminergic reward circuitry, is widely implicated in the pathophysiology of depression^32,33^ and mediates positive valence-related emotional response and reward processing^4,31,34–39^. Furthermore, functional connectivity (FC) between the NAc and prefrontal cortex is shown to relate to anhedonia in depression^35,38^ and antidepressant response^40^.

Another key region involved in reward behavior that may be relevant to the therapeutic mechanisms of ketamine is the habenula (Hb) located near the posterior and medial region of the thalamus^41^. The lateral component of the habenula (LHb) projects to dopaminergic VTA and serotonergic raphe nucleus, where it inhibits monoaminergic activity, therefore regulating reward behavior^42,43^ and is often referred to as the ‘anti-reward’ system^42,44^. Preclinical and human studies strongly implicate LHb dysfunction as contributing to depression and anhedonia, and suggest its involvement in antidepressant response^42,43,45–47^. Furthermore, the blockade of LHb NMDAR bursting activity is specifically linked with the rapid antidepressant action of ketamine in preclinical models^48,49^, supporting the hypothesis that ketamine’s therapeutic effects involve modulation of reward-related networks. In humans, a recent resting state fMRI study (rsfMRI) reported that increased FC between the Hb and frontal pole, occipital pole/cortex, temporal pole and parahippocampal gyrus associated with antidepressant response following single-dose ketamine^50^.

The literature supports the role of the NAc and Hb in reward dysfunction in MDD and suggests that ketamine may target these deficits. To further understand how ketamine modulates antidepressant and reward-related brain circuitry, the current investigation addressed how FC of the NAc and Hb to other brain regions are modulated by ketamine at the brain systems level. Although a majority of rsfMRI studies have looked at FC by investigating a fixed relationship between brain regions over multiple minutes, FC is not necessarily static. Rather, the connectivity between brain regions experience fluctuations over short time periods, which has motivated the investigation of dynamic FC^51–53^. Dynamic FC investigations can be used to measure temporal variations of connectivity strength, and disruptions in dynamic FC have been implicated in various psychiatric disorders including MDD^54–57^. Therefore, in addition to examining changes in static FC (sFC), dynamic FC was investigated by measuring dynamic FC variability (dFCv), defined as the standard deviation of FC across a series of windowed timeseries throughout fMRI acquisition. To accomplish this goal, advanced rsfMRI acquisition and computational analysis methods were combined to determine whether Hb or NAc to whole brain FC change over time and associate with improvements in mood and anhedonia in TRD participants receiving serial ketamine infusion (SKI) treatment. Based on the limited prior literature^50,58^, we hypothesized that changes in NAc and Hb FC with frontal, temporal and occipital regions would change over time and associate with improved mood or anhedonia.

## Methods and Materials

### Participants

Fifty-eight depressed participants (average age=40.7 years, 28 female) participated in this naturalistic clinical trial (NCT02165449) (https://clinicaltrials.gov/study/NCT02165449). Participants received four serial intravenous ketamine infusions over the course of 2 weeks. MRI scanning and clinical and behavioral data were acquired 1) prior to treatment (baseline), occurring <1 week before the first ketamine infusion, and 2) 24 hours after the last ketamine infusion (**Figure 1**).

**Figure 1.**
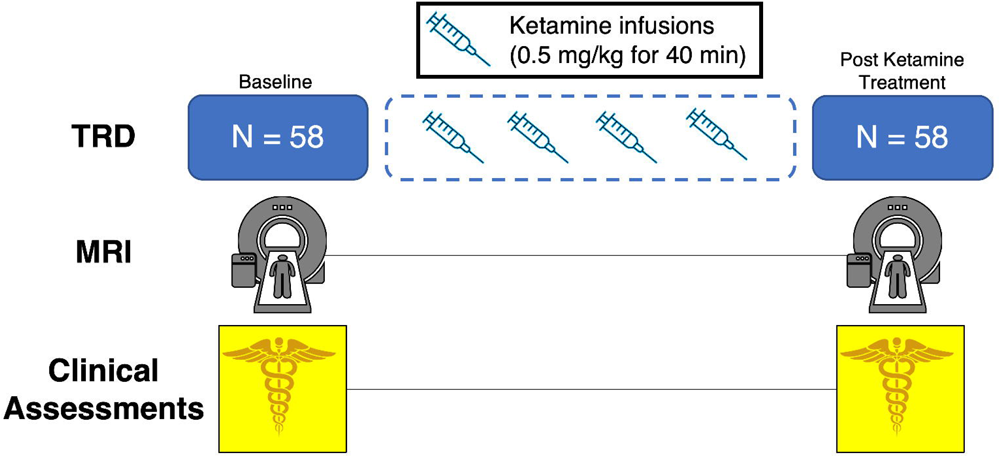
Overview of study design. Treatment Resistant Depression (TRD) patients received MRI scans (T1w, T2w, resting-state fMRI) at baseline, and 24 to 72 hours after receiving 4 intravenous ketamine infusions, over a 2 week period.

Participants were included if they met criteria for TRD, which was defined as nonresponse to ≥2 adequate antidepressant trials of adequate dosage and duration, and being continuously depressed for ≥6 months. Additional eligibility criteria for participants included DSM-V-defined major depression^59^, moderate to severe depressive symptoms as per the Hamilton Depression Rating Scale (HDRS) 17-item (scores ≥ 17)^60^, no prior psychotic reactions to medications, alcoholic or illicit substances in the past, or other physical or clinical contraindications to ketamine. Exclusion criteria included any unstable medical or neurological condition, current substance abuse or dependence (measured by laboratory testing) or substance abuse history within the preceding 3-months, current or past history of psychosis, schizophrenia, intellectual disability or other developmental disorder, diagnosis of dementia and any contraindication to scanning (e.g., metal implants or claustrophobia). Subjects were recruited from the Los Angeles area through advertisements, clinician referral or clinicaltrials.gov. All subjects provided written informed consent following procedures approved by the UCLA Institutional Review Board (IRB).

### Ketamine Treatment

Ketamine treatment was administered 2-3 times a week and included intravenous pump delivery of a sub-anesthetic dose (0.5 mg/kg) of racemic ketamine diluted in 60cc normal saline. Participants were permitted to remain on approved monoaminergic antidepressant therapy if dosage was unchanged in the preceding 6-weeks. Benzodiazepines were discontinued >72 hours prior to all study visits including scanning sessions.

### Clinical Assessments

At each time point, depression severity was assessed using the Hamilton Depression Rating Scale (HDRS)^60^, and anhedonia was measured with the Snaith-Hamilton Pleasure Scale (SHAPS)^61^. Remitters were defined as Participants with a HDRS score of ≤7 post-SKI treatment.

### Image Acquisition

Imaging was performed on a Siemens 3T Prisma MRI system at the UCLA Brain Mapping Center using a 32-channel head coil. Imaging sequences were identical to those used by the Human Connectome Project (HCP) Lifespan studies for Aging and Development^62^. Structural sequences included T1-weighted (voxel size (VS)=0.8mm isotropic; repetition time (TR)=2500ms; echo time (TE)=1.81:1.79:7.18ms; inversion time (TI)=1000ms; flip angle (34)=8.0°; acquisition time (TA)=8:22min), and T2-weighted acquisitions (VS=0.8mm isotropic; TR=3200ms; TE=564ms; TA=6:35min), both with real-time motion correction^63^. Resting state fMRI used a multi-band EPI sequence with opposite phase encoding directions over two runs (VS=2mm isotropic; TR=800ms; TE=37ms, FA=52°, MB accl. factor=8; phase enc. direction=AP(run1)/PA (run2); TA=13:22 min), along with two sets of spin echo images used for distortion correction^64,65^. During rsfMRI, subjects viewed a fixation cross.

### Image Processing and Denoising

Imaging data was preprocessed using the HCP minimal processing pipelines^66^. Processing of rsfMRI data included ICA-based denoising using FSL’s multi-run FIX (https://fsl.fmrib.ox.ac.uk/fsl/fslwiki/FIX), and alignment using MSMAll^67^. Further denoising was performed using the CONN Toolbox^68^, which included component-based noise correction (CompCor) and band-pass filtering. A band-pass filter of [0.015 -0.1] Hz was used to remove high-frequency noise and low-frequency activity with a period exceeding the duration of sliding windows while still preserving relevant low frequency signal, as done in prior studies^51,54,69^. Specific frequency ranges were used since prior research suggests that the window length for sliding window analyses should be no less than 1/*f_min_* (the minimum frequency in the timeseries, defined by the band-pass filter) to prevent spurious correlations^51^. Images were then converted to CIFTI space with 4mm smoothing using the grayordinates-based approach^70^. The quality of the functional data was assessed using relative and absolute motion plots. All subjects moved less than 3mm, therefore, no subjects were excluded.

### Hb and NAc Seed Generation

For each subject and timepoint, the right and left habenula were segmented using validated methods^71,72^. To generate more reliable segmentations and to obtain a single segmentation mask for each subject per hemisphere, the high-resolution segmented Hb maps were averaged across timepoints for each subject. Signals from 3mm spheres located in adjacent thalamic nuclei were regressed out from each Hb seed to prevent signal contamination^71^. The Harvard-Oxford atlas (https://fsl.fmrib.ox.ac.uk/fsl/fslwiki/Atlases) was used to obtain left and right NAc masks in standard space. Using the segmented and atlas defined Hb and NAc masks, the average timeseries was extracted from the denoised images for all subjects and timepoints to generate Hb and NAc seeds.

### Resting-State Connectivity Maps

The average Hb and NAc timeseries were used to generate seed-based sFC and dFCv maps in CIFTI space. Seed-based sFC maps were generated using HCP workbench commands^73^ (https://www.humanconnectome.org/software/workbench-command) to calculate the correlation between the timeseries from the seeds and every other vertex and voxel in the brain, followed by fisher-z transformation.

Seed-based dFCv maps were calculated using the sliding-windows analysis approach^51,74,75^, implemented using in-house code developed in Python (https://github.com/btaraku/SeedBased_dynamic_FC). Window length is an important parameter, since sliding windows should be short enough to identify resting-state fMRI fluctuations, while also being long enough to avoid detecting spurious correlations. We opted to follow similar temporal filtering and window lengths to another study using high resolution HCP fMRI data^69^. The higher temporal resolution fMRI used in this study enables us to sample a larger number of TRs within a shorter window of time compared to prior dFCv studies^55,56,76^, since more data points decrease the influence of noise on the correlation estimates^51^. Here, we used a window length of 84 TRs (approximately 67 seconds) since this is no less than 1/*f_min_*^51^. Additionally, we used a Hamming window, since tapered window functions may limit detecting outlier points near window boundaries^51,53,56^, and used a step size of 1 TR (0.8 seconds). Correlations were computed across all vertices and voxels within each window and were followed by fisher-z transformation. Once correlations were generated for all windows, the sample standard deviation was calculated across windows to calculate dFCv, which were then converted back to CIFTI grayordinate space. dFCv results were validated by trying different window lengths (125 TRs (100 seconds) and 63 TRs (50s)). **Figure 2** provides a flow chart of the imaging processing and fMRI analysis steps used to generate seed-based sFC and dFCv maps in CIFTI space.

**Figure 2.**
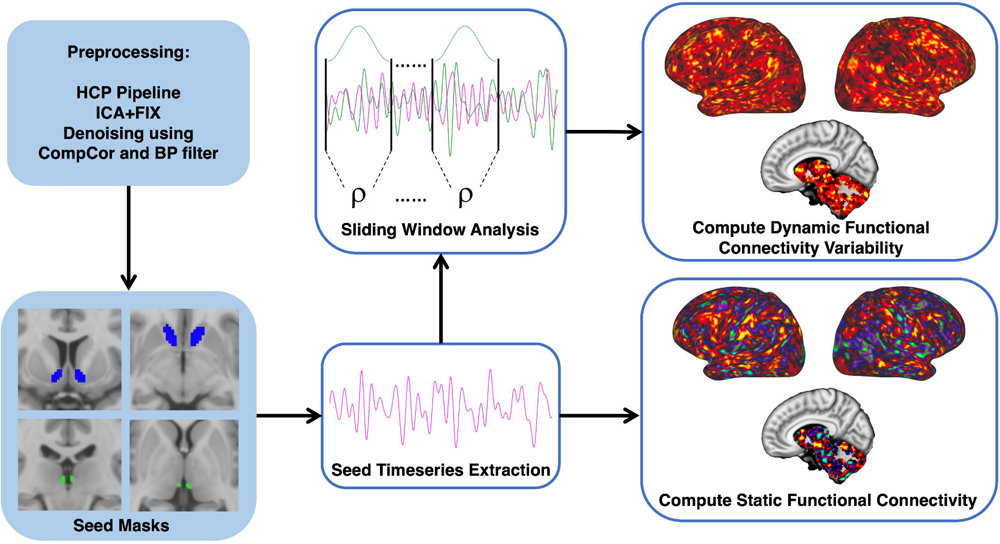
Overview of fMRI processing. Resting-state fMRI was preprocessed using HCP protocols, which includes motion correction, correcting for EPI induced distortions and the conversion to CIFTI grayordinate space, and was followed by further denoising in the CONN toolbox. Habenula masks were acquired for each subject following an automated segmentation protocol using T1w and T2w images, while the Nucleus Accumbens masks were obtained from the Harvard-Oxford atlas in standard space and were used for all subjects. The average fMRI timeseries was extracted from each seed mask and used to generate whole-brain functional connectivity (FC) maps. Static FC was computed directly from the extracted timeseries, whereas dynamic FC variability was computed after running a sliding window analysis.

### Statistical Analyses

To examine ketamine’s effect on our clinical measures of interest, paired t-tests were performed to determine whether HDRS or SHAPS scores significantly changed in participants following SKI.

Voxelwise group level analyses were conducted in PALM^77^ (https://fsl.fmrib.ox.ac.uk/fsl/fslwiki/PALM) and designed to test two primary hypotheses: 1) effects over time pre-to-post SKI in participants, and 2) associations between change in FC and change in mood and anhedonia. All analyses were run using 5000 randomly generated permutations, using Threshold-Free Cluster Enhancement (TFCE) and Family-wise error rate correction (FWER). Age and sex were included as covariates of no interest in all analyses.

To test for longitudinal changes following SKI, the baseline sFC and dFCv maps were subtracted from the post-treatment sFC and dFCv maps to compute change across time for each subject, which were then used to run a 1-sample t-test in PALM.

To test whether sFC or dFCv changes were significantly associated with improvements in depressive symptoms and anhedonia, correlations between change in sFC and dFCv and percent change in each clinical score (HDRS, SHAPS) was calculated at every vertex/voxel using PALM. To prevent spurious correlations, outliers with extreme values (+/-3 standard deviations from the mean) were removed prior to running PALM. As a result, one subject was removed from the HDRS correlation analysis and one subject was removed from the SHAPS correlation analysis. One additional subject was removed from both analyses due to missing clinical data. For visualization of effects, the average change was extracted from significantly correlated brain regions (TFCE/FWER corrected) and used to map associations with changes in mood on a scatter plot. Post-hoc correlations were calculated in MATLAB using partial correlations controlling for age and sex.

## Results

### Demographic and Clinical Results

Participants with TRD showed significant improvements in both HDRS (t=-13.496, p=4.3e-19), and SHAPS (t=7.64, p=3.27e-10) scores following SKI treatment. Of the 58 participants that completed SKI, 29 reached remission status. **Table 1** shows an overview of participant demographics and symptom changes over time following SKI.

**Table 1.** Demographics and clinical values by group and time point.

|  | MDD Mean (SD) |  |  |  |
| --- | --- | --- | --- | --- |
|  | Baseline | Post-SKI | t-score | p-value |
| HDRS | 19 (4.76) | 8.4 (4.6) | -13.49 | 4.30E-19 |
| SHAPS | 32.35 (6.79) | 39.95 (7.02) | 7.64 | 3.27E-10 |
| Number of subjects (N) | 58 |  | - | - |
| Sex (% female) | 56.36 |  | - | - |
| Age (years) | 40.7 (11.3) |  | - | - |
| Education (ISCED variable) | 5.79 (1.2) |  | - | - |
| Duration of lifetime illness (years) | 24.78 (16.3) |  | - | - |
| Current episode (years) | 8.89 (21.98) |  | - | - |
| Race (% Asian) | 10.53 |  | - | - |
| Race (% Black) | 0.00 |  | - | - |
| Race (% Hawaiian or Pacific Islander) | 0.00 |  | - | - |
| Race (% more than one race) | 1.75 |  | - | - |
| Race (% unknown/not reported) | 7.02 |  | - | - |
| Race (% White) | 80.70 |  | - | - |

Abbreviations: MDD: major depressive disorder; SD: standard deviation; HDRS: Hamilton Depression Rating Scale; SHAPS: Snaith-Hamilton Pleasure Scale

### Effect of treatment on Hb and NAc FC

Significant increases in sFC were found between the left Hb and bilateral visual cortex, as well as significant decreases in sFC between the left Hb and left inferior parietal cortex. Significant decreases in sFC were found between the left NAc and right cerebellum (all p<0.05, TFCE/FWER corrected) (**Figure 3**). No significant changes in dFCv were observed over time.

**Figure 3.**
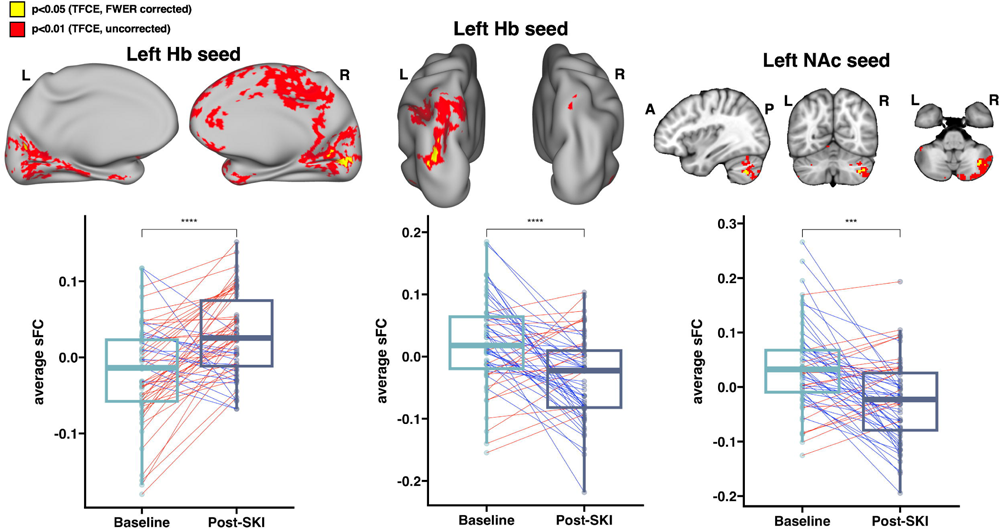
Longitudinal effects of ketamine treatment on functional connectivity. Paired t-tests comparing FC change before and after ketamine treatment revealed **(A)** significant increases in static FC between the left Habenula (Hb) and bilateral visual cortex, **(B)** significant decreases in static FC between the HbL and left inferior parietal cortex, and **(C)** significant decreases in static FC between the left Nucleus Accumbens (NAc) and right cerebellum (all p<0.05, FWER and TFCE corrected). Boxplots show average static FC before and after ketamine treatment averaged within significant regions across all treatment resistant depression (TRD) participants. Brain images include results in surface and voxel space, and display corrected p-values overlaid on top of uncorrected p-values, in order to show trends. *Acronyms: (sFC: static Functional Connectivity, Hb: Habenula, NAc: Nucleus Accumbens, SKI: Serial Ketamine Infusions, TFCE: Threshold-Free Cluster Enhancement, FWER: Family-wise error rate)*

### Associations between changes in FC and changes in clinical symptoms

Decreases in dFCv between the left Hb, right precuneus and right visual cortex, as well as between the right NAc and right visual cortex were both significantly correlated with improvements in HDRS following SKI (**Figure 4**). Results of this analysis using different window lengths can be seen in **Supplemental Figure 1**. Decreases in sFC between the left Hb and bilateral visual and parietal cortex, right precuneus, right angular gyrus, right fusiform gyrus and right somatomotor cortex were significantly correlated with improvements in SHAPS following SKI. Increases in sFC between the left NAc and left visual cortex, left parietal cortex, and right fusiform gyrus were significantly correlated with improvements in SHAPS following SKI (**Figure 5**) (all p<0.05, TFCE/FWER corrected).

**Figure 4.**
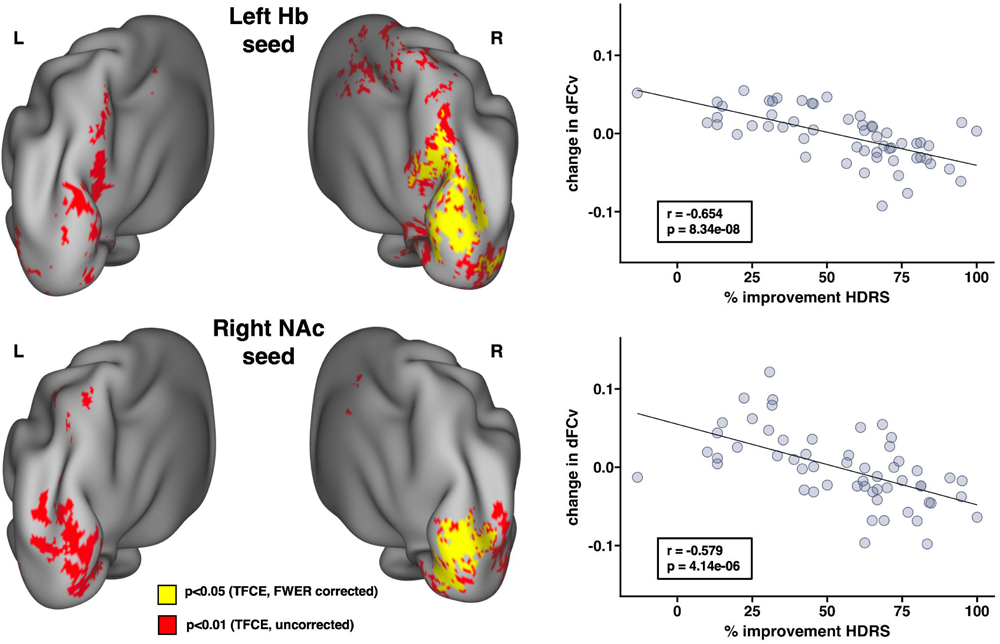
Associations between changes in dynamic functional connectivity variability (dFCv) and improvements in depressive symptoms following ketamine treatment. Whole brain correlations were performed to investigate how FC changes were associated with improvements in HDRS scores. Results revealed that decreases in dFCv between **(A)** the left Habenula (Hb) and right precuneus and visual cortex, as well as decreases in dFCv between the **(B)** right Nucleus Accumbens (NAc) and right visual cortex were significantly associated with improvements in HDRS (all p<0.05, FWER and TFCE corrected). Overlapping effects between the Hb and NAc were observed within a region of the right visual cortex. Average dFCv change was calculated within significant regions to create scatter plots between dFCv and HDRS change, in order to visualize the overall effects within these regions. R and P values are shown on each scatter plot, and brain images show corrected p-values overlaid on top of uncorrected p-values to display trending effects. *Acronyms: (FC: functional connectivity dFCv: dynamic FC variability, Hb: Habenula, NAc: Nucleus Accumbens, TFCE: Threshold-Free Cluster Enhancement, FWER: Family-wise error rate)*

**Figure 5.**
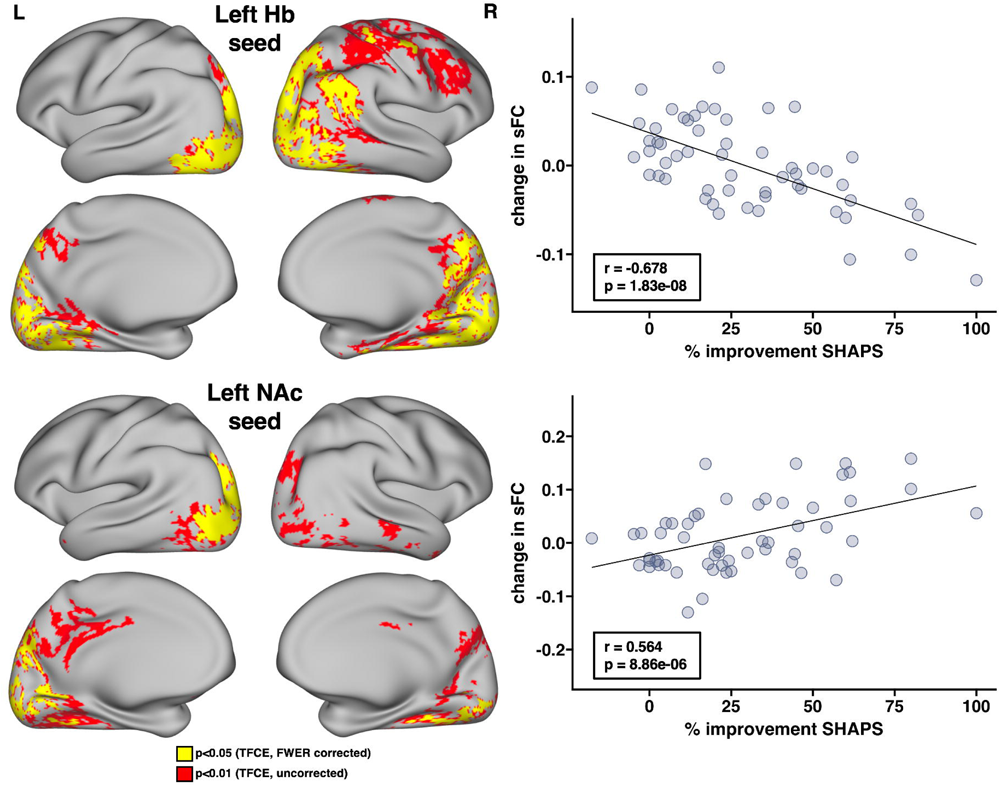
Associations between changes in static functional connectivity (sFC) and improvements in anhedonia following ketamine treatment. Whole brain correlations were performed to investigate how FC changes were associated with improvements in anhedonic symptoms using the SHAPS. Results revealed that decreases in sFC between **(A)** the left habenula (Hb) and bilateral visual and parietal cortex as well as the right precuneus, right temporoparietal junction, right fusiform gyrus and right somatomotor cortex were significantly associated with improvements in SHAPS. Additionally, increases in sFC between **(B)** the left nucleus accumbens (NAc) and left visual and parietal cortex, as well as the right fusiform gyrus were significantly associated with improvements in SHAPS (all p<0.05, FWER and TFCE corrected). Overlapping effects between the Hb and NAc were observed within regions of the left visual and parietal cortex, as well as the right fusiform gyrus. Average sFC change was calculated within significant regions to create scatter plots between sFC and SHAPS change to visualize the pattern of effects within these regions. R and P values are shown on each scatter plot, and brain images show corrected p-values overlaid on top of uncorrected p-values to display trending effects. *Acronyms: (FC: functional connectivity sFC: static FC, Hb: Habenula, NAc: Nucleus Accumbens, TFCE: Threshold-Free Cluster Enhancement, FWER: Family-wise error rate)*

## Discussion

Recent evidence suggests that sub-anesthetic ketamine may modulate the brain’s reward systems to reduce depressive symptoms and anhedonia^19–22^. In this study, we used advanced image analysis methods to study how functional connectivity of brain networks involving the Hb and NAc, key nodes of reward circuitry previously implicated in the neurobiology of depression^45,48,78^, are perturbed after SKI treatment in participants with TRD, and to investigate whether FC changes relate to improvements in mood and anhedonia. To address these questions, we acquired high resolution HCP resting-state fMRI data and used state-of-the-art preprocessing pipelines, followed by seed-to-whole brain analyses to examine static and dynamic functional connectivity.

We observed significant changes in sFC following SKI that included increases between left Hb and bilateral visual cortex, decreases between left Hb and left posterior parietal cortex, as well as decreases in sFC between the left NAc and right cerebellum. Furthermore, reduced dFCv between the left Hb and contralateral precuneus and visual cortex and reduced dFCv between right NAc and visual cortex were found to be significantly associated with antidepressant response. In addition, increases in sFC between the left Hb and bilateral parietal and visual regions, and reductions in sFC between left NAc and left parietal and visual regions were found to be significantly associated with improvements in anhedonia specifically. Together these findings suggest ketamine’s therapeutic effects occur via overlap in the functional circuitry of the Hb and NAc.

### Effect of treatment on Hb and NAc FC

Significant increases in sFC between the Hb and the visual cortex were observed over the course of SKI treatment in TRD participants. To date, the majority of published FC studies of depression have focused on examining differences in large-scale functional networks such as the DMN, salience (SN) and fronto-parietal/central executive (FPN/CEN) and limbic networks. However, alterations in visual networks have also been associated with depression^79,80^. Further, three studies from our own group employing different imaging modalities and including an overlapping TRD sample have reported ketamine-related modulation of visual areas, including changes in BOLD activation during a response inhibition task^81^, increases in cerebral blood flow^82^, and changes in white matter microstructure within occipital white matter pathways^83^. Notably, one of the few published studies investigating the effects of ketamine on habenula FC found that increases in sFC between the habenula and occipital pole and lateral visual cortex following a single ketamine infusion significantly correlated with improvements in subjectively rated depressive symptoms^50^. Another prior study found that MDD participants exhibited decreased long range functional connectivity density in several regions of the visual cortex compared to controls, highlighting that the visual cortex may be an important network hub for MDD treatment^84^. These findings combined with those of the current study suggest that changes in subcortical and visual networks contribute to ketamine’s antidepressant effects.

We also observed decreases in sFC between Hb and inferior parietal lobule (IPL) following ketamine treatment in TRD participants. The IPL is contained within the posterior parietal cortex (PPC)^85^, which forms a key node within the frontoparietal network (FPN). The FPN is involved in the top-down regulation of attention and emotion and nodes within the FPN generally show hypoconnectivity in depression^86^. Thus, perturbation of Hb-FPN circuitry by ketamine may contribute to its antidepressant effects.

Decreases in sFC between the nucleus accumbens and cerebellum were also observed pre-to-post ketamine in TRD participants. Although the cerebellum is classically viewed as involved in motor coordination, much evidence shows the cerebellum also contributes to cognitive and emotional function^87,88^. fMRI studies suggest the cerebellum and basal ganglia are strongly interconnected, forming a network involved in reward and learning^89^. Several studies have found functional abnormalities in cerebellar circuitry in depressed participants^90–92^. In addition, we have previously found that ketamine treatment induces functional changes in the cerebellum during a response inhibition task-fMRI^93^. The current findings thus further implicate cerebellar-striatal circuitry as a target for antidepressant treatments.

### Associations with clinical response

Since longitudinal changes may be the consequence of biological effects independent of antidepressant effects, we examined whether changes in Hb and NAc circuitry associated with improvements in depressive symptoms and anhedonia. We found that changes in both sFC and dFCv were associated with improvements in overall mood and anhedonia. While sFC measures temporal correlations and can determine whether there is a strong or weak excitatory or inhibitory relationship between brain regions, dFCv determines the degree of fluctuations in connectivity strength between brain regions, which are also reported as relevant to psychiatric disease states^54–56,94–97^. Prior studies have reported differences in dynamic FC from the Hb^95^ and striatum^56,57^ in depressed participants when compared to healthy controls, further highlighting dynamic alterations in reward circuitry as a potential biomarker for MDD.

Our findings show that greater decreases in dFCv between Hb, precuneus and visual cortex, as well as between NAc and visual cortex, are associated with greater treatment response to ketamine. Although we also found that ketamine treatment significantly increases sFC between Hb and visual cortex, these findings together do not necessarily conflict. Rather, decreases in dFCv implies less temporal variability in connectivity strength, which suggests a convergence towards a stable connectivity value within these networks as depressive symptoms improve. Furthermore, these two findings could be considered complementary, since we observed a small overlap between these two regions, and it is possible that the observed increases in Hb-visual cortex FC could lead to downstream effects that result in reductions of FC variability across broader regions of the visual cortex.

Although several previous studies have investigated dynamic FC alterations in MDD within reward circuitry, no previous studies to our knowledge have investigated how dynamic FC is altered in Hb or NAc networks in MDD following ketamine treatment. One study investigating ketamine’s effect on dynamic FC in non-depressed healthy controls using whole brain atlases found that ketamine produces an overall negative effect on dynamic FC within visual networks^98^, which complement our results. Since we observed associations in overlapping regions of the left visual cortex for both Hb and NAc seeds, this suggests dynamic visual cortex activity may contribute to reward circuitry, and ketamine’s antidepressant effects could stabilize activity between these visual and reward networks. Further research is necessary to understand the precise biological mechanism driving the interplay between visual and reward systems that may underlie ketamine’s antidepressant effects.

Improvements in depressive symptoms were also correlated with changes in dFCv between the Hb and precuneus. The precuneus is a key node of the DMN, which has been widely implicated in major depression^99–101^, and is often associated with self-referential thinking and ruminative symptoms of depression^102,103^. Dynamic FC disruptions in the DMN have been reported in MDD, including studies that have found decreases in dFCv within core DMN components^54,104^, and increases in dFCv between DMN and other brain regions including dlPFC and insula, which also correlated with symptom severity^54^ in depressed individuals compared with controls. However, another study reported increased dFCv within the core DMN components in depressed participants compared to controls^105^. Of relevance to this study, greater dFCv between the habenula and right precuneus was found in depressed participants with suicide ideation compared to healthy controls, although opposite trends were found for the contralateral precuneus^95^. Interestingly, the precuneus is also suggested to be involved in the dissociative effects induced by ketamine, where reductions in oscillations measured by electroencephalography in the precuneus are associated with dissociation^106,107^. Thus, it may be the case that ketamine’s antidepressant effects and dissociative effects are driven by similar neural mechanisms. In conjunction with prior findings, our results suggest that ketamine’s antidepressant effects may modulate dynamic FC disruptions between the Hb and precuneus.

Decreases in sFC between Hb and posterior parietal and visual cortex, angular gyrus, precuneus, motor cortex, and fusiform gyrus, as well as increases in sFC between NAc and overlapping regions in the visual cortex, parietal cortex and fusiform gyrus were associated with improvements in anhedonia. The NAc forms an integral part of the brain’s reward circuitry and shows hypoactivity in depression, whereas the Hb is part of the antireward system and shows hyperactivity in depression. Our results suggest the anti-anhedonic effects of ketamine are driven by perturbations of aberrant reward circuitry; increasing NAc and subsequently decreasing Hb connectivity in broadly distributed overlapping and distinct brain regions. The overlapping regions occurred primarily in the visual cortex, posterior parietal cortex and fusiform gyrus. Relevant to our findings, a prior study found reduced FC between NAc and IPL in participants with depression and also found a significant interaction between anhedonia and diagnosis on FC between NAc and IPL^35^, highlighting that parietal regions may contribute to anhedonic symptoms.

In addition to visuo-parietal regions, significant decreases in FC between Hb and the precuneus and angular gyrus, key nodes of the DMN, were correlated with improvements in anhedonia. Of relevance to this finding, a prior study reported hyperconnectivity between the habenula and precuneus which was associated with suicidality in TRD^47^. Thus, our results suggest ketamine normalizes hyperconnectivity between Hb and DMN. Decreases in Hb and somatomotor cortex FC were also found to be significantly associated with improvements in anhedonia. Prior imaging studies have reported altered FC of somatosensory and somatomotor networks correlate with depressive symptoms, including suicidality^108,109^. Preclinical studies also suggest Hb inhibition of dopamine neurons leads to suppression of body movements and to lowered motivation^110^.

### Limitations

We acknowledge that there are several limitations associated with the current investigation. Firstly, the Hb is a small structure, and the spatial resolution of imaging protocols may still be inadequate for fully capturing structural and functional changes in this region even when using advanced acquisition and preprocessing methods. We also note that the current study was not a randomized clinical trial and did not include an active control condition. However, we note that this was designed as a mechanistic clinical trial focused on imaging outcomes rather than on the efficacy of SKI. Also, participants were allowed to continue concurrent stable antidepressant medication, which may have impacted findings although we note that participants serve as their own controls in longitudinal analyses and did not change medication throughout the trial.

### Conclusion

Serial ketamine treatment in TRD modulates static and dynamic FC in both overlapping and distinct Hb and NAc functional networks. SKI produces distinct static FC changes from Hb and NAc seeds in TRD participants, whereas static and dynamic FC changes associated with clinical response occurred in broader overlapping regions from Hb and NAc seeds, suggesting that antidepressant response to SKI involves reward, sensory and self-referential networks.

## Supporting information

Suuplemental Figure 1

## Data Availability

The data that support the findings of this study are publicly available in the NIMH Data Archive in collection 2844

https://nda.nih.gov/edit_collection.html?id=2844

## Acknowledgements

This work was supported by the National Institute of Mental Health of the National Institutes of Health (Grant Nos. <u>MH110008 [to KLN and RE]</u>, and <u>MH102743 [to KLN</u>]). The content is solely the responsibility of the authors and does not necessarily represent the official views of the National Institutes of Health.

