## Supplementary material for "Ketamine treatment modulates habenular and nucleus accumbens static and dynamic functional connectivity in major depression": Suuplemental Figure 1

**Supplemental Information**

**Effect of Window Size**

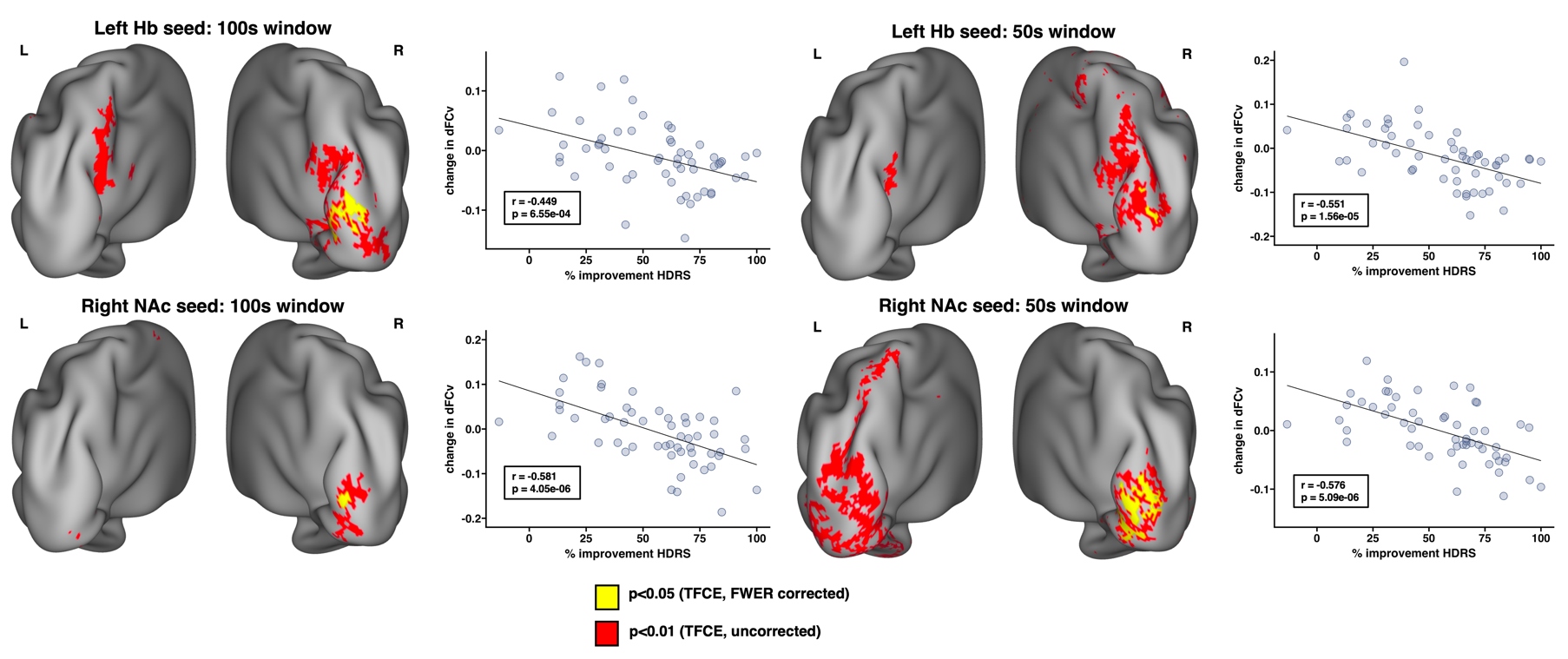

**Supplemental Figure 1:** Associations between changes in dFCv and HDRS after modifying window length. A sliding hamming window with a length of 125 TRs (100s) and 63 TRs (50s) were used to repeat the analysis investigating correlations between change in HDRS and change in dFCv, in order to examine the effect of window length on our results. In addition to modifying the window length, we reprocessed our data using a 0.01 Hz high-pass filter for the 100s window and a 0.02 Hz high-pass filter for the 50s window, as suggested by Leonardi and Van De Ville et al^1^. All other parameters and preprocessing steps remained the same as described in the Methods section. Results remain similar to those seen in **Figure 4** which show significant clusters in the right visual cortex, however, the strongest effects were observed with an 84 TR (67s) window, which may suggest this window size is more optimal for capturing rfMRI fluctuations.
